# Transtibial amputation increases the metabolic energy needed for stabilizing walking in the sagittal plane

**DOI:** 10.1101/2025.09.12.25335253

**Authors:** Wouter Muijres, Maarten Afschrift, Renaud Ronsse, Friedl De Groote

## Abstract

Walking energy consumption is higher in people with versus without a transtibial amputation, but the underlying reasons are poorly understood. Active prostheses that restore ankle power fail to decrease walking energy consumption, suggesting that there should be other reasons than a lack of ankle power. Transtibial amputation impacts walking stability, as evidenced by the increased fall risk, and there is an energetic cost associated with stabilizing walking. It is, however, unclear how transtibial amputation affects the energetic cost of stabilizing walking. We assessed the metabolic cost of stabilizing walking against treadmill belt speed perturbations (SD=0.13 m/s) in 16 subjects with and 23 subjects without a transtibial amputation at three walking speeds between 0.6 and 1.6 m/s. We focused on sagittal plane stability, as a transtibial amputation mainly affects modulation of the ankle torque, and this ankle strategy contributes most to sagittal plane control of walking at low speeds. Perturbations induced 0.24 W/kg larger increases in energy consumption across walking speeds in subjects with than without a transtibial amputation. Whereas mean reductions in step length in response to perturbations were similar between groups, individuals with an amputation increased step length variability of their intact leg more – especially at low speeds – than individuals without an amputation. As continuous control is required for unperturbed walking, an increased metabolic cost of stabilizing walking might explain – at least partially - the higher energetic cost of walking. These insights are important when seeking to reduce the metabolic cost of walking after transtibial amputation.

**NEW & NOTEWORTHY:** The higher metabolic cost of walking in individuals with versus without a transtibial amputation is poorly understood, hindering the design of interventions. We demonstrated that transtibial amputation considerably increases the energy consumption for stabilizing walking in the sagittal plane and increases the reliance on step length adjustments to stabilize walking. This opens up perspectives for restoring walking energetics after amputation through prostheses that support sagittal plane stability.

## INTRODUCTION

People with a transtibial amputation using a prosthesis consume on average 12% (nonvascular amputation) to 36% (vascular amputation) more metabolic energy to walk than individuals without an amputation (1) but the underlying reasons are not well understood. A lack of ankle power is thought to be an important reason for higher energy consumption (2). Yet active prostheses that restore ankle power fail to decrease walking energy consumption (3). This suggests that a lack of ankle power may not be the main cause for the higher walking energy consumption in individuals with a transtibial amputation. Transtibial amputation also severely impacts walking stability as it results in increased fall risk (4). It has been shown that there is an energetic cost associated with stabilizing walking (5–9) but it is unclear how this is affected by transtibial amputation. Transtibial amputation is expected to affect sagittal plane balance control. Individuals without an amputation use an ankle strategy, i.e., they adjust the center of pressure (COP) under the foot through modulation of ankle muscle activation, to stabilize walking against sagittal plane perturbations (10) but this capacity is lost in people using prosthetic ankles. Transtibial amputation is thus expected to affect sagittal plane balance control. Individuals with a transtibial amputation may rely more on stepping strategies, i.e., anticipatory or reactive adjustments in step length, and this might come at a metabolic cost. In this paper, we assessed the metabolic cost of stabilizing walking against sagittal plane perturbations in individuals with and without a unilateral transtibial amputation and its association with the strategy used to stabilize walking.

### Although ankle power has been related to the metabolic cost of walking in healthy individuals, restoring ankle power has only yielded limited decreases in the metabolic cost of walking in individuals with a transtibial amputation

In healthy individuals, an ankle range of motion constraining orthosis that decreased push-off power by 50% increased energy consumption by up to 50% (11). This suggests that the ability to generate ankle power contributes considerably to walking energetics in individuals without an amputation. Indeed, augmentation of ankle power using an exoskeleton decreased walking energy consumption by 24% in healthy adults without an amputation (12). Most prostheses are passive and – in contrast to biological ankles - do not generate net positive mechanical work, which might be a reason for the higher metabolic cost of walking in individuals with an amputation. However, bionic ankle prostheses that restore ankle power in individuals with a transtibial amputation fail at normalizing walking energy consumption in most individuals with a transtibial amputation (3, 13, 14), even when the prosthesis generated high push-off power (3). While ankle power is an important contributor to walking energetics in individuals without an amputation, restoring ankle power in individuals with a transtibial amputation does not yield the expected reductions in metabolic energy consumption. This suggests that there might be other reasons for the higher metabolic cost of walking in individuals with an amputation.

### There is an energetic cost associated with stabilizing walking but it is unclear whether this cost differs between individuals with and without a transtibial amputation

In individuals without an amputation, external lateral stabilization of the pelvis reduces energy consumption by 5.7% (6) while external perturbations during walking increase walking energy consumption by 28% for unstable surfaces (5), 5.9-12% for mediolateral visual field perturbations (7, 8), and 16.7% for sagittal plane speed belt perturbations (9). Together, these studies demonstrate that there is a considerable energetic cost associated with stabilizing walking in both the frontal and sagittal planes. In individuals with an amputation, handrail support decreased the energy consumption of walking on a treadmill by on average 11% (15). Given the small forces on the handrail, the decrease in energy consumption was attributed to the stabilizing effect of handrail support. However, it is unclear whether the effect of handrail support on metabolic cost is different or similar than in individuals without an amputation. Lateral stabilization of the pelvis had a similar effect on walking energy consumption in individuals with and without a transtibial amputation (16). An important limitation of this study was that pelvis stabilization impeded gait compensations that are often observed in individuals with an amputation and the increase in metabolic cost due to alterations in the gait pattern caused by constraining pelvis motion might have masked a larger decrease in the energetic cost of stabilizing walking in individuals with an amputation (15, 16). It remains unclear whether the metabolic cost of stabilizing walking is increased in individuals with versus without a transtibial amputation, as these two studies investigating frontal plane stabilization are inconclusive and sagittal plane stabilization has not yet been investigated in individuals with an amputation.

### In healthy adults without an amputation, the energetic cost to stabilize walking is associated with the control strategy used to stabilize walking

Healthy individuals use a combination of anticipatory and reactive strategies to stabilize walking. They decrease mean step length and step width when expecting a perturbation (17, 18), these adaptations in gait kinematics can thus be considered as anticipatory strategies. They also modulate step length (5, 9) and width (7, 8) as well as their ankle torque (10) in response to perturbations. These modulations have been associated with deviations in center of mass kinematics and can thus be considered as reactive strategies (10, 19). The extent to which each of these strategies is used depends on the condition and differs between individuals. For example, an ankle strategy is more effective for stabilizing walking in the sagittal plane than in the frontal plane as the dimensions of the foot allow for larger COP displacements in the fore-aft than in the mediolateral direction (10). Individuals who increased step width and step width variability (reflecting ongoing modulation of step width) more in response to frontal plane perturbations and decreased mean step length more in response to sagittal plane perturbations, increased metabolic cost more (7, 9). Given these associations between the control strategy and metabolic cost of stabilizing walking, alterations in control strategies due to amputation might induce alterations in the metabolic cost.

### Whereas the inability to use an ankle strategy on the prosthetic side might mainly affect sagittal plane stabilization of walking, the focus of previous studies was on frontal plane balance control in individuals with an amputation

Individuals with versus without a transtibial amputation walk with wider steps irrespective of whether walking is perturbed (20). Individuals with a transtibial amputation increase step width variability more, suggesting that they rely more on reactive stepping, in response to mediolateral platform perturbations (21), unstable surfaces (22, 23), and frontal plane swing foot perturbations (24) than individuals without an amputation. Alterations in sagittal plane balance control strategies have not been investigated, possibly because walking is thought to be more robust against small perturbations in the sagittal than in the frontal plane (25). However, the loss of the ankle strategy on the amputated side likely affects the ability to stabilize walking in the sagittal plane. Indeed, slips and trips, which result in backward and forward balance loss, are the most common causes of falls among individuals with a lower limb amputation (26). The loss of the ankle strategy is expected to induce a larger reliance on anticipatory and reactive stepping adjustments in step length, which might have consequences for the metabolic cost of stabilizing walking, but this remains to be investigated.

### In this study, we investigated whether individuals with a unilateral transtibial amputation used different strategies and more metabolic energy to stabilize walking against sagittal plane perturbations than individuals without amputation

To this aim, we applied continuous treadmill belt speed perturbations and assessed step length and metabolic energy use. We previously found that belt speed perturbations elicit ankle and stepping responses, and increase the energy cost of walking in healthy adults without amputation but more so at slow than fast walking speeds (9, 10). We hypothesized that individuals with a transtibial amputation rely more on stepping responses, i.e. decreases in mean step length and increases in step length variability, due to the inability to use an ankle strategy on the amputated side. Since anticipatory reductions in step length have been associated with an energetic cost (9), we hypothesized that individuals with versus without a transtibial amputation would consume more energy to stabilize walking against treadmill belt speed perturbations. Furthermore, we explored relationships between strategies and energy consumption for stabilizing walking in individuals with and without an amputation.

## METHODS

We recruited 16 individuals with a unilateral transtibial amputation in the age range of 19-76 years old (see Table 1 for an overview of subject characteristics). In addition, we used previously collected data of 23 healthy subjects without an amputation between 19-76 years old for comparison (9). All subjects provided written informed consent. Individuals were excluded from participation if they suffered from conditions or took medication that could influence balance control and locomotor abilities (other than amputation) or were unable to walk for at least 5 minutes without walking aid, pain, or socket discomfort. Transtibial amputation due to traumatic etiology was most common in our study sample (i.e., 13/16 subjects). We evaluated mobility in subjects with a transtibial amputation using the PLUS-M 12-Item Short Form - version 1.2 (27). The mean t-score on the PLUS-M Short Form was on average 59.1 (see Table 1), which is higher than at least 50% of individuals with a traumatic unilateral below-knee amputation included in the PLUS-M reference dataset (t-score = 55.4) (28). The experimental protocol was approved by the Ethics Committee Research UZ / KU Leuven (S69186) and pre-registered on Open Science Framework (29) after collecting data from subjects without an amputation and the first subject with an amputation.

**Table 1.** Summary table of subject group characteristics, with in the left column the group of subjects without an amputation and in the right column the group of subjects with a unilateral transtibial amputation. Standard deviation (sd) is displayed between parentheses for age in years, body mass in kilograms, leg length in meters, and the PLUS-M score.

| Group | Without amputation | With unilateral transtibial amputation |
| --- | --- | --- |
| N | 23 | 16 |
| Age (years) | 41.3 (sd=15.8) | 58.7 (sd=12.9) |
| Sex | 12 female | 3 female |
| Body mass (kg) | 71.8 (sd=15.5) | 83.7 (sd=13.8) |
| Leg length (m) | 0.93 (sd=0.06) | 0.93 (sd=0.04) |
| Etiology | NA | 3 vascular |
| Side amputation | NA | 6 right |
| PLUS-M score | NA | 59.1 (sd=7.4) |

### Experimental procedure

All subjects walked with standardized shoes and subjects with amputation used their prescribed prosthesis on an instrumented treadmill (M-Gait, Motekforce link, Amsterdam, Netherlands) at three different speeds with and without treadmill belt speed perturbations. We aimed for subjects with a transtibial amputation to walk at 0.8, 1.2, and 1.6 m/s to match the dataset previously collected in individuals without an amputation (9) but adapted these speeds to accommodate individuals’ walking ability and to ensure safety. In a habituation trial, subjects with a transtibial amputation walked at different speeds, with and without perturbations. The experimenter increased walking speed starting from 0.8 m/s with increments of 0.2 m/s and let the subject experience walking with and without perturbations. After 1 minute, subjects were asked whether they thought they would be able to walk 5 minutes in the experienced condition. Based on verbal feedback on the different walking conditions, the experimenters selected a slow, medium, and fast speed out of six walking speeds (i.e., 0.6, 0.8, 1.0, 1.2, 1.4, and 1.6 m/s). In both subjects with and without an amputation, walking conditions were randomized per block, i.e., first by speed and then by perturbation condition (Figure 1A). In each trial, subjects walked for about 6 minutes with at least 5 minutes of rest between walking trials. During walking trials, subjects wore a safety harness, tethered to the ceiling, to prevent falls in the event of balance loss. Not all subjects with a transtibial amputation completed all six walking trials. Two subjects completed 4 of the 6 trials because they were unable to complete the last walking conditions within the time of the experiment (+/-1.5 hours) due to prolonged periods of rest between trials, or because of the time needed to address equipment failure. In another subject, motion capture data in one of the trials was lost due to equipment failure. Finally, yet another subject was able to complete all 6 walking trials but only completed 3-4 of the 5 minutes of walking in perturbed walking trials due to fatigue. We decided to include this data and estimated metabolic rate based on the last two minutes (as in other trials). The shorter duration of the trials might have led to an underestimation of the metabolic rate as it takes about 3 minutes for metabolic rate estimates to reach steady-state (30) and therefore including these trials would have decreased rather than increased differences between individuals with and without an amputation.

**Figure 1.**
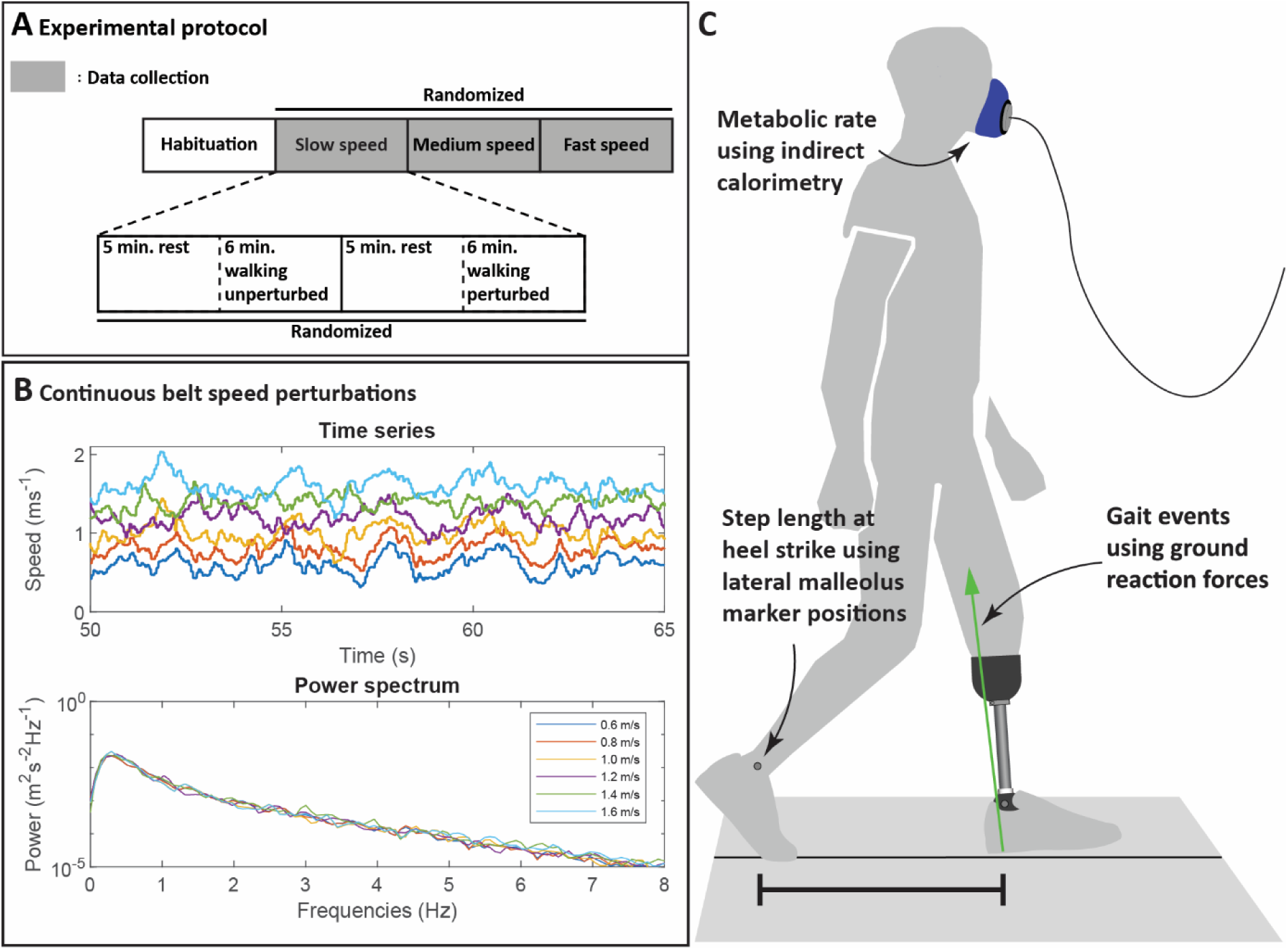
Summary of the experimental protocol. **A.** Schematic representation of the order of experimental conditions and the randomization procedure. **B.** The time series of the observed treadmill belt velocity in perturbation trials measured at 300Hz (top plot), and the power spectrum (bottom plot) showing the frequency content of the measured treadmill belt velocities for the six different speeds for representative subjects. **C.** Visualization of measured data.

### Data acquisition

We measured oxygen consumption and carbon dioxide production to estimate metabolic energy consumption using the K5 metabolic measurement unit (COSMED, Rome, Italy). Subjects were asked not to consume caffeine (12 hours), nicotine (12 hours), or food (3 hours) before the experiment to prevent variability in measured signals unrelated to physical effort. Marker and ground reaction force (GRF) data were acquired through the Vicon Giganet system (Oxford Metrics, Oxford, England) in combination with a Vicon camera system to measure the position of both lateral malleoli or, in the case of subjects with an amputation, a marker on the lateral side of the prosthesis at the height of the intact leg’s malleolus using retroreflective markers at 200 Hz to estimate step length at heel strike. Gait events were detected based on GRF, which were measured on the M-Gait instrumented treadmill at 1000 Hz.

### Perturbation protocol

During perturbation trials, walking was perturbed by continuous random fore-aft treadmill belt speed variations. To construct the belt speed control signal for the perturbation trials, we adapted the procedure described in (31). The control signals for the velocity-controlled treadmill were generated in MATLAB and Simulink using software that was distributed with (31). First, we created pseudo-random belt accelerations using discrete Gaussian white noise with a standard deviation of 5 m/s^2^ and bounded between -15 and 15 m/s^2^. Then, we integrated the accelerations to generate a velocity perturbation signal. The velocity perturbation signal was high-pass filtered at 1.7 Hz to eliminate drift, and added to a constant speed of either 0.6, 0.8, 1.0, 1.2, 1.4, or 1.6 m/s, depending on the condition. The resultant speeds were bounded between 0 and 3.6 m/s. We kept the perturbation magnitude constant over walking speeds. In contrast to (31), the velocity perturbation signal for all walking speeds was generated using the same level of random noise, acceleration bounds, and high-pass filter cut-off frequency. We measured treadmill belt speed variation using the treadmill belt speed sensors. The standard deviation of measured velocities was approximately 0.13 m/s. Perturbations did not affect the intended average belt speed, and velocity bounds were never reached (Figure 1B). Measured treadmill belt velocities during perturbed walking at different speeds had similar frequency contents, indicating a similar perturbation profile across walking speeds.

### Data pre-processing

GRF and marker data were collected using Vicon’s Nexus. We labeled marker data in Nexus and processed data in MATLAB (R2023, MathWorks, Natick, USA). GRF and marker data were low-pass filtered using a third-order bidirectional Butterworth filter with a cut-off frequency of 20 Hz. Periods of missing marker data shorter than 0.1 seconds were linearly interpolated to account for marker occlusion.

Heel strike and toe-off were defined as the moment at which the vertical component of the GRF under the respective foot increased above or decreased below 10% of body weight, respectively. Gait events could not be accurately determined when both feet were in contact with the same belt of the split-belt treadmill. To eliminate steps with both feet on the same belt, we first assessed the distance between the foot and the edge of the belt. When the marker on the big toe, first metatarsal head, or medial malleolus (or the equivalent position on the prosthesis) was within 4 cm of the edge of the belt, we only included the step if additional criteria guaranteeing single foot contact were met (see Supplementary material S6).

### Outcomes

#### Metabolic energy consumption

We estimated metabolic rate from oxygen consumption and carbon dioxide production, measured by the K5 device, using the standard formula of Brockway (17). The energy consumption of the walking trials was represented by the average metabolic rate over the last two minutes of the trial.

#### Step length

Step length was estimated by calculating the fore-aft distance between markers on the lateral malleoli at heel strike of the leading leg. We used the difference in mean step length to evaluate anticipatory changes to the gait pattern, and the difference in standard deviation of step length to evaluate changes in reactive foot placement. The mean step length and variability were calculated over the last 3 minutes of walking, as a trade-off between including enough steps in the analysis to obtain reliable outcomes and overlap with the period over which the metabolic rate was determined.

We normalized metabolic rate to g^3/2^l^1/2^m, and step length to l to obtain all outcomes in dimensionless units (i.e. n.d.), where g is the gravity constant, l is leg length measured between the right lateral malleolus and posterior superior iliac spine, and m is the body mass of the subject. In subjects with a prosthesis, body mass included the weight of the prosthesis. Normalization factors for a subject with an average leg length and body mass were 2272 W for metabolic rate, and 0.93 m for step length.

### Statistical analysis

The study design of this experiment is mixed with a between-subject dichotomous variable, group (with and without an amputation), and a within-subject continuous variable, speed. Statistical analysis was performed using R statistical software (R Foundation for Statistical Computing, Vienna, Austria). For testing our hypotheses, we used the difference in outcome variables between perturbed and unperturbed conditions as dependent variables in statistical models. We evaluated the effect of group and speed by fitting separate linear mixed-effect models on the change in metabolic rate, anticipatory stepping (mean step length), and reactive stepping (standard deviation in step length) outcomes to test for the effect of walking speed with a fixed and random slope for speed and a random intercept per subject.

We added the factor Leg to linear mixed models in which we assessed changes in mean step length and step length variability, which could be different for both legs. The factor Leg accounted for the difference between the prosthetic and intact leg in individuals with a transtibial amputation and between the left and right leg for individuals without an amputation. We chose to separate the step outcomes for the left and right leg to reduce the number of linear mixed-effect models that were needed and to balance the reliability of the stepping measures equally. If we had averaged step outcomes over the left and right legs of individuals without an amputation, this would have resulted in different levels of precision (i.e., degree of variance with repeated measurement) in step length outcomes between subjects with and without an amputation. We expected that separating step length outcomes for both legs in both groups would lead to more balanced variances over groups, which aids the statistical comparison.

In addition, we explored associations between increases in metabolic cost and changes in kinematic strategies (mean step length and step length variability) and the influence of transtibial amputation on these associations using linear mixed-effect models. Here, we included speed, group, leg, and balance outcomes as fixed effects, a random offset per subject, and a random slope with speed. We accounted for the interaction between speed and balance outcomes.

Linear mixed-effect models were reduced to their simplest form through stepwise elimination based on F-tests for marginal effects and likelihood ratio tests for random effects. In case the reduced model contained interaction effects between group and speed, we performed a post-hoc analysis. We first predicted outcomes from the linear mixed-effect models for 0.8, 1.2, and 1.6 m/s and then tested for differences at 0.8, 1.2, and 1.6 m/s. We used R^2^ of the fixed effects to quantify the effect sizes. We corrected for multiple comparisons to prevent Type-I error inflation due to the number of statistical tests that we performed using the Benjamini-Hochberg procedure. Corrections for Type-I error inflation were performed over confirmatory hypotheses, and separately for the different exploratory hypotheses. We considered probability values lower than 0.05 as statistically significant.

### Departures from preregistration

Hypotheses and methods were preregistered before conducting this study (29). We decided to deviate from the preregistered document on different points. The most important departures from the pre-registration include the decision to move the analysis of step width to the supplementary material; include post-hoc comparisons to test for differences between groups in the effect of perturbations on metabolic and stepping outcomes per speed; to accommodate for multiple comparisons using the Benjamini-Hochberg procedure; and to include separate stepping measures for both legs in the linear-mixed effect models, i.e. left and right in subjects without an amputation, and intact and prosthetic in subjects with a transtibial amputation. A more elaborate description of the changes with respect to the pre-registered document can be found in the supplementary material S1.

## RESULTS

Statistical tests were performed after scaling outcomes to dimensionless units. For the sake of interpretability, we report in the text the corresponding values for the average subject, i.e., a subject with a leg length of 0.93 m and a body mass of 76.7 kg. Corresponding non-dimensional values can be found between parentheses and in Tables 2-4.

**Table 2.** Fixed effects in linear mixed-effect models that predict metabolic rate during unperturbed walking (Eĳ_unperturbed_), change in metabolic rate due to perturbations (Eĳ), change in mean step length due to perturbations (ΔMean step length), and change in step length variability due to perturbations (ΔStep length var.) based on speed, group (without or with amputation) and leg (left or right for individuals without amputation; intact or prosthesis for individuals with amputation). The Predictors column presents the effects after the full model was step-wise reduced. Group effects represent differences in subjects with compared to those without a transtibial amputation in model estimates. TAMP for the group effects indicates changes in estimates in subjects of the group with amputation compared to subjects without an amputation. I or L for the leg effect indicates differences in estimates for the intact or left with respect to the prosthetic or right leg. R²-values represent the fit of the fixed effects with the data. The Estimates (Scaled) column displays model estimates that were scaled by mean body mass and leg length, the Estimates (CI) column shows model estimates and 95% confidence intervals based on data that was normalized to body mass and leg length, and the p column shows probability values, which were corrected by the Benjamini-Hochberg procedure.

| Model | Predictors | Estimate (Scaled) | Estimates (CI) | $p$ |
| --- | --- | --- | --- | --- |
| $\dot{E}_{\text{unperturbed}}$<br>($R^2 = 0.72$) | Intercept | -46 W | -0.020 (-0.031 - -0.009) | < 0.01 |
|  | Speed | 251 Ws/m | 0.111 (0.101 - 0.120)<br>s/m | < 0.001 |
|  | Group [TAMP] | 44 W | 0.020 (0.009 - 0.030) | < 0.01 |
|  | Intercept | 70 W | 0.031 (0.021 - 0.040) | < 0.001 |
| $\Delta \dot{E}$<br>($R^2 = 0.14$) | Speed | -28 Ws/m | -0.012 (-0.019 - -0.005)<br>s/m | < 0.01 |
|  | Group [TAMP] | 19 W | 0.008 (0.001 - 0.016) | 0.05 |
| ΔMean step length<br>($R^2 = 0.03$) | Intercept | -0.039 m | -0.042 (-0.053 - -0.031) | < 0.001 |
|  | Speed | 0.012 s | 0.013 (0.005 - 0.021) | < 0.01 |
| ΔStep length var.<br>($R^2 = 0.61$) | Intercept | 0.052 m | 0.056 (0.049 - 0.062) | < 0.001 |
|  | Speed | -0.023 s | -0.025 (-0.030 - -0.020)<br>s/m | < 0.001 |
|  | Group [TAMP] | 0.006 m | 0.006 (-0.004 - 0.017) | 0.26 |
|  | Leg [I/L] | -0.003 m | -0.003 (-0.012 - 0.005) | 0.49 |
|  | Speed – group [TAMP]<br>interaction | -0.007 s | -0.007 (-0.016 - 0.001)<br>s/m | 0.12 |
|  | Speed – leg [I/L]<br>interaction | 0.001 s | 0.001 (-0.006 - 0.008)<br>s/m | 0.72 |
|  | Group [TAMP] – leg [I/L]<br>interaction | 0.024 m | 0.026 (0.012 - 0.04) | 0.001 |
|  | Speed – group [TAMP] –<br>leg [I/L] interaction | -0.013 s | -0.014 (-0.025 - -0.002)<br>s/m | 0.04 |

**Table 3.** Post-hoc results for step length variability in which statistically significant interaction between group and speed were found. Test results are organized first by outcome and then by walking speed. The comparison column describes what comparison is made. TAMP (resp. NAMP) refers to the subjects with (resp. without) an amputation. In adaptations in step length variability, an interaction effect between group and leg was found. Therefore, comparisons for the left [L], right [R], intact [I], and prosthetic [P] legs were separated. Differences between groups scaled to average leg length and body mass are reported in the Difference (Scaled) column, and normalized to the subjects’ body mass and leg length in the Difference column. The SE column presents the standard errors (SE) that were estimated from the linear mixed-effect models, and the t-statistic column shows the t-ratio that was calculated based on the difference and the SE. P-values obtained from the statistical analysis were corrected for the number of comparisons using the Benjamini-Hochberg correction. The last column shows the effects size using Cohens’ d, for which absolute values of <0.2 indicate small, 0.2 - 0.5 medium, and > 0.8 large effect sizes.

| Outcome | Speed | Comparison | Difference<br>(Scaled) | Difference | SE | t-statistic | P | Cohens'<br>d |
| --- | --- | --- | --- | --- | --- | --- | --- | --- |
| ΔStep length var. | 0.8<br>m/s | NAMP [R] -<br>TAMP [P] | -0.001 m | -0.001 | 0.003 | -0.23 | 1.00 | -0.09 |
|  |  | NAMP [R] -<br>NAMP [L] | 0.002 m | 0.002 | 0.002 | 1.22 | 1.00 | 0.33 |
|  |  | NAMP [R] -<br>TAMP [I] | -0.012 m | -0.013 | 0.003 | -5.21 | <<br>0.001 | -1.98 |
|  |  | TAMP [P] -<br>NAMP [L] | 0.003 m | 0.003 | 0.003 | 1.10 | 1.00 | 0.42 |
|  |  | TAMP [P] -<br>TAMP [I] | -0.012 m | -0.013 | 0.002 | -6.48 | <<br>0.001 | -1.89 |
|  |  | NAMP [L] -<br>TAMP [I] | -0.015 m | -0.016 | 0.003 | -6.08 | <<br>0.001 | -2.31 |
|  | 1.2<br>m/s | NAMP [R] -<br>TAMP [P] | 0.002 m | 0.002 | 0.002 | 1.07 | 1.00 | 0.35 |
|  |  | NAMP [R] -<br>NAMP [L] | 0.002 m | 0.002 | 0.001 | 1.50 | 0.99 | 0.26 |
|  |  | NAMP [R] -<br>TAMP [I] | -0.005 m | -0.006 | 0.002 | -2.50 | 0.21 | -0.82 |
|  |  | TAMP [P] -<br>NAMP [L] | -0.001 m | -0.001 | 0.002 | -0.27 | 1.00 | -0.09 |
|  |  | TAMP [P] -<br>TAMP [I] | -0.007 m | -0.008 | 0.002 | -4.97 | <<br>0.001 | -1.16 |
|  |  | NAMP [L] -<br>TAMP [I] | -0.007 m | -0.007 | 0.002 | -3.29 | 0.03 | -1.07 |
|  | 1.6<br>m/s | NAMP [R] -<br>TAMP [P] | 0.005 m | 0.005 | 0.003 | 1.71 | 0.83 | 0.78 |
|  |  | NAMP [R] -<br>NAMP [L] | 0.001 m | 0.001 | 0.002 | 0.68 | 1.00 | 0.19 |
|  |  | NAMP [R] -<br>TAMP [I] | 0.002 m | 0.002 | 0.003 | 0.76 | 1.00 | 0.35 |
|  |  | TAMP [P] -<br>NAMP [L] | -0.004 m | -0.004 | 0.003 | -1.31 | 1.00 | -0.60 |
|  |  | TAMP [P] -<br>TAMP [I] | -0.003 m | -0.003 | 0.003 | -0.98 | 1.00 | -0.43 |
|  |  | NAMP [L] -<br>TAMP [I] | 0.001 m | 0.001 | 0.003 | 0.36 | 1.00 | 0.16 |

**Table 4.** Prediction of between-subject variability in changes in metabolic rate based on between-subject variability in either changes in mean step length or step length variability due to perturbations, speed and group. The model column indicates the different models that were fitted to predict the change in metabolic rate. The predictor column contains the model terms after step-wise reduction of the full model. The effect of groups indicates the change in metabolic rate in subjects who belonged to the groups with a transtibial amputation (TAMP). The Estimates (Scaled) column shows model estimates scaled to a subject with average body mass and leg length, and the Estimates (CI) column shows the estimates and 95% confidence intervals normalized to body mass and leg length. The p column presents the probability values of estimates after correction using the Benjamini-Hochberg procedure.

| Model | Predictors | Estimates<br>(Scaled) | Estimates (CI) | p |
| --- | --- | --- | --- | --- |
| ΔMean step length<br>($R^2 = 0.19$) | Intercept | 34 W | 0.015 (0.010 – 0.020) | p<0.001 |
| | ΔMean step length | -1235 W/m | -0.505 (-0.639 – -0.371) | p<0.001 |
|  | Group [TAMP] | 17 W | 0.008 (0.000 – 0.015) | p=0.06 |
| | ΔMean step length - speed<br>interaction | 938 Ws/m<sup>2</sup> | 0.384 (0.268 – 0.499) s/m | p<0.001 |
| ΔStep length var.<br>($R^2 = 0.15$) | Intercept | 13 W | 0.006 (-0.000 – 0.012) | p=0.07 |
| | ΔStep length var. | 1030 W/m | 0.421 (0.254 – 0.589) | p<0.001 |
|  | Group [TAMP] | 35 W | 0.015 (0.005 – 0.025) | p<0.01 |
| | ΔStep length var. – group [TAMP] interaction | -656 W/m | -0.269 (-0.503 – -0.034) | p=0.03 |

The metabolic rate in unperturbed walking trials increased with walking speed and was higher in subjects with compared to those without an amputation, irrespective of walking speed (Figure 2A). Metabolic rate of unperturbed walking increased by 251 W for every 1 m/s (0.111 s/m, CI=0.101 - 0.120 s/m, p < 0.001) increase in walking speed (Table 2). An average subject without an amputation consumed 159 W at 0.8 m/s, 258 W at 1.2 m/s, and 357 W at 1.6 m/s (Table S1). The metabolic rate of unperturbed walking was 44 W higher (0.020, CI=0.009 – 0.030, p<0.01) in subjects with compared to those without an amputation (Table 2). Subjects with a transtibial amputation consumed 198 W at 0.8 m/s, 298 W at 1.2 m/s, and 397 W at 1.6 m/s (Table S1), which is respectively 25%, 15%, and 11% more than individuals without an amputation.

**Figure 2.**
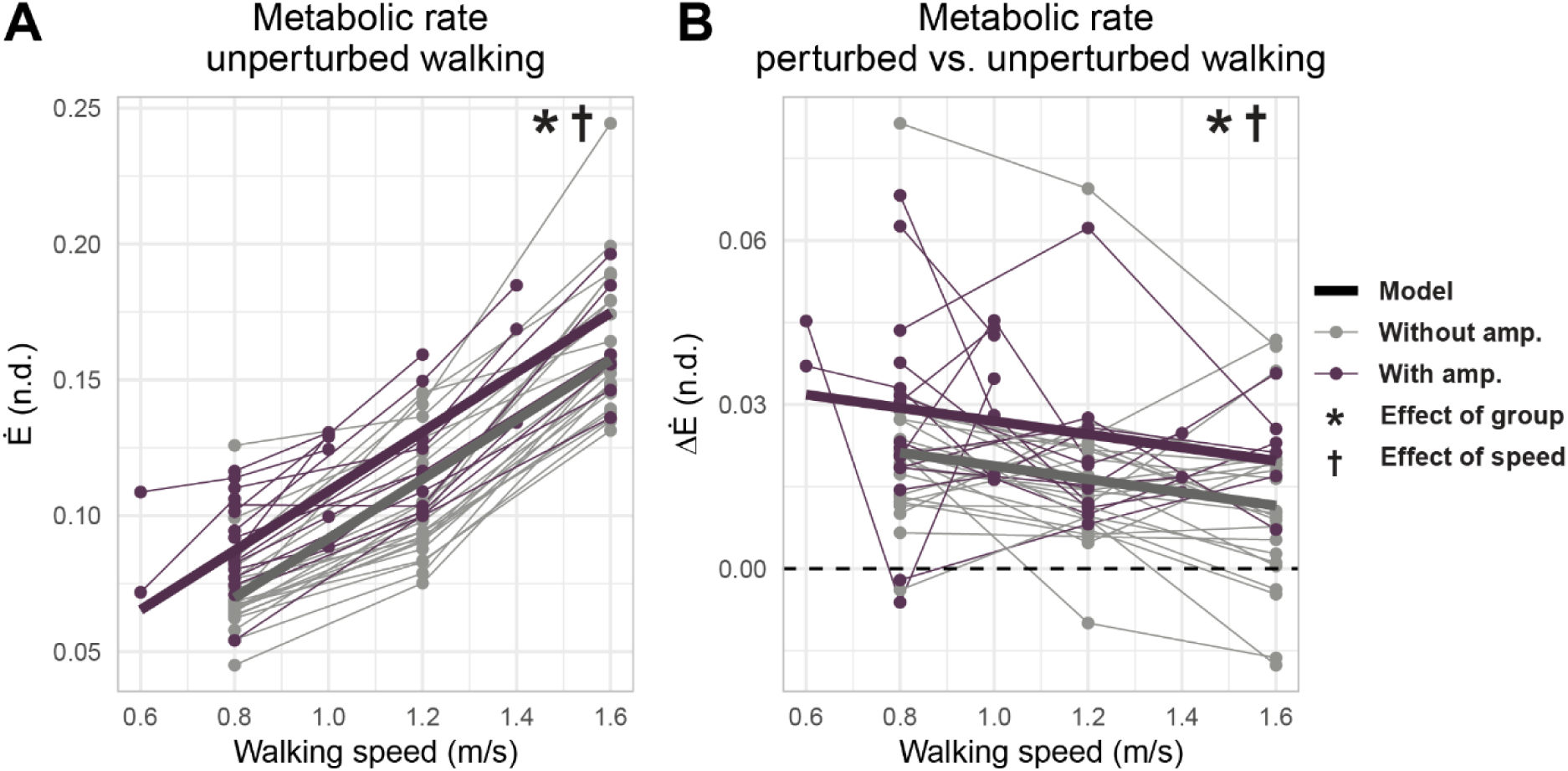
A. Normalized metabolic rate in individuals with (purple) and without (grey) a transtibial amputation during unperturbed walking at different walking speeds. **B.** Differences in metabolic rate between perturbed and unperturbed walking for individuals with and without an amputation. Dots and connecting lines depict measured data, while the thick lines represent the fixed effects obtained through fitting linear mixed-effect models. * denotes a significant effect of group and ^†^ a significant effect of walking speed.

Perturbations increased the energy cost of walking more at slow than at fast walking speed in all individuals, but individuals with a transtibial amputation consumed more energy and by a similar amount at each speed (Figure 2B). The energy needed to stabilize walking against perturbations decreased by 28 W with every 1 m/s (0.012 s/m, CI=-0.019 – -0.005 s/m, p < 0.01) of increase in walking speed (Table 2). As a result, the average subject without an amputation consumed 48 W to stabilize walking at 0.8 m/s, 37 W at 1.2 m/s, and 26 W at 1.6 m/s. The energy needed to stabilize walking was 19 W (0.008, CI=0.001 - 0.016, p=0.04) higher in subjects with a transtibial amputation and independent of walking speed. Subjects with a transtibial amputation consumed 67 W at 0.8 m/s, 56 W at 1.2 m/s, and 45 W at 1.6 m/s to stabilize walking against perturbations.

Individuals reduced mean step length in response to balance perturbations, and changes in mean step length decreased with increasing walking speed, similarly in individuals without and with an amputation (Figure 3A). With every m/s increase in walking speed, subjects shortened steps by 0.01 m less (0.013 s/m, CI=0.005 - 0.021 s/m, p < 0.01). Subjects shortened mean step length in response to perturbations by 0.030 m at 0.8 m/s, 0.025 m at 1.2 m/s, and 0.020 m at 1.6 m/s (Table S1).

**Figure 3.**
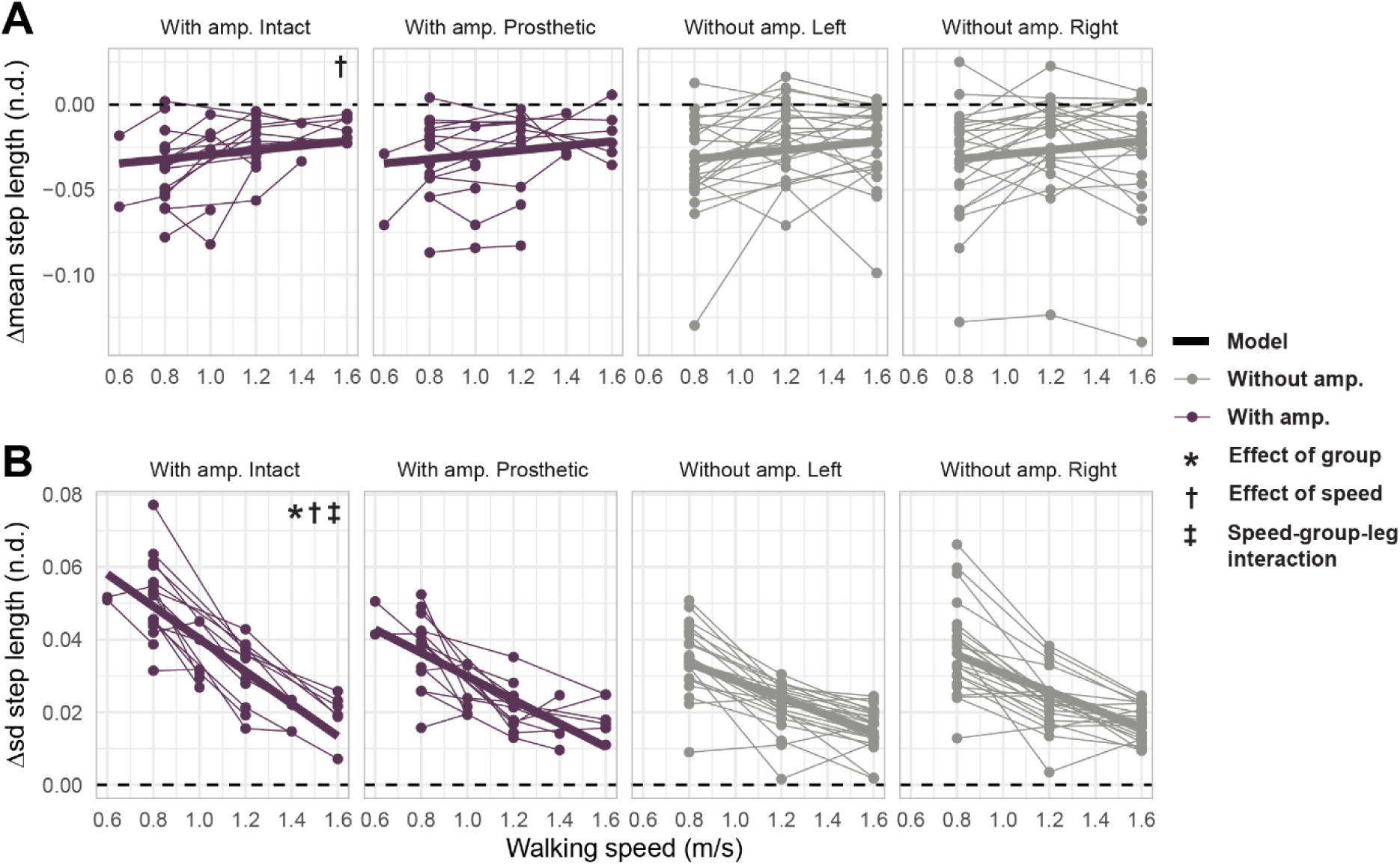
Changes in mean step length (A) and standard deviation (sd) of step length (B) in response to treadmill belt speed perturbations for the intact (first column) and prosthetic (second column) legs of subjects with an amputation (amp.), and left (third column) and right (fourth column) legs of subjects without an amputation. Data from each individual subject is presented by dots, connected through thin lines. The thick lines represent fixed effects derived from the mixed-effects models. Statistically significant effects of the linear mixed effect models are depicted in the first column of each row. * denotes a significant effect of group, ^†^ a significant effect of walking speed, and ^‡^ a significant speed-group-leg interaction effect.

Step length became more variable in response to perturbations but the increase in variability depended on walking speed and group with the largest increases at the lowest speed for the intact leg of individuals with a transtibial amputation (Figure 3B). Subjects without an amputation decreased step length variability by 0.020 m less for every 1 m/s (-0.024 s/m, CI=-0.029 - -0.019 s/m, p < 0.001) increase in walking speed for both the left and right leg (Table 2). In the left leg, subjects without an amputation increased step length by 0.031 m at 0.8 m/s, 0.022 m at 1.2 m/s, and 0.013 m at 1.6 m/s, and in the right leg by 0.033 m at 0.8 m/s, 0.024 m at 1.2 m/s, and 0.015 m at 1.6 m/s. Compared to individuals without an amputation, step length variability was 0.030 m (group-leg interaction term: 0.026, CI=0.012 – 0.040, p<0.01) larger in the intact leg of subjects with a transtibial amputation (Table 2). Additionally, with every 1 m/s increase in walking speed, adaptations in step length variability decreased by 0.019 m (speed-group-leg interaction term: -0.014 s/m, CI=-0.025 - -0.002 s/m, p=0.04) more in the intact leg of subjects with an amputation compared to subjects without an amputation (Table 2). Subjects with a transtibial amputation increased step length variability in response to perturbations on the intact side by 0.046 m at 0.8 m/s, 0.029 m at 1.2 m/s, and 0.013 m at 1.6 m/s, and on the prosthetic side by 0.034 m at 0.8 m/s, 0.022 m at 1.2 m/s, and 0.011 m at 1.6 m/s (Table S1). At a walking speed of 0.8 m/s, step length variability of the intact leg of subjects with a transtibial amputation was 0.012 m larger (p < 0.001) than the prosthetic leg, 0.012 m larger (p < 0.001) than the right leg, and 0.015 m larger (p < 0.001) than the left leg of subjects without an amputation (Table 3). At a walking speed of 1.2 m/s, step length variability of the intact leg of subjects with a transtibial amputation was 0.7 cm larger than the prosthetic leg (p < 0.001) and the left leg of subjects without an amputation (p = 0.03). At a walking speed of 1.6m/s, post-hoc testing revealed no differences in step length variability between legs (Table 3).

We explored whether inter-subject differences in increases in metabolic rate in response to perturbations could be explained by inter-subject differences in changes in mean step length and step length variability (Figure 4). Our exploratory analysis indicates that increases in metabolic rate in response to perturbations can in part be explained by adaptations in mean step length (R^2^=0.19) and step length variability (R^2^=0.15). Subjects who decreased mean step length more in response to perturbations increased their metabolic rate more (Figure 4A). A decrease in step length of 1 cm increased metabolic rate by 4.8 W (or 484.11 W/m) when walking at 0.8 m/s, but metabolic rate increased by 9.4 W/cm (or 938 W/m) less for every 1 m/s increase in walking speed (Table 4). Additionally, metabolic rate increased more in subjects whose steps were more variable in response to perturbations, but less so in subjects with compared to without an amputation (Figure 4B). While subjects without an amputation increase metabolic rate by 10.3 W for every cm increase in step length variability (or 1030 W/m), individuals with a transtibial amputation increased metabolic rate by 6.6 W/cm (or 656 W/m) less (Table 4).

**Figure 4.**
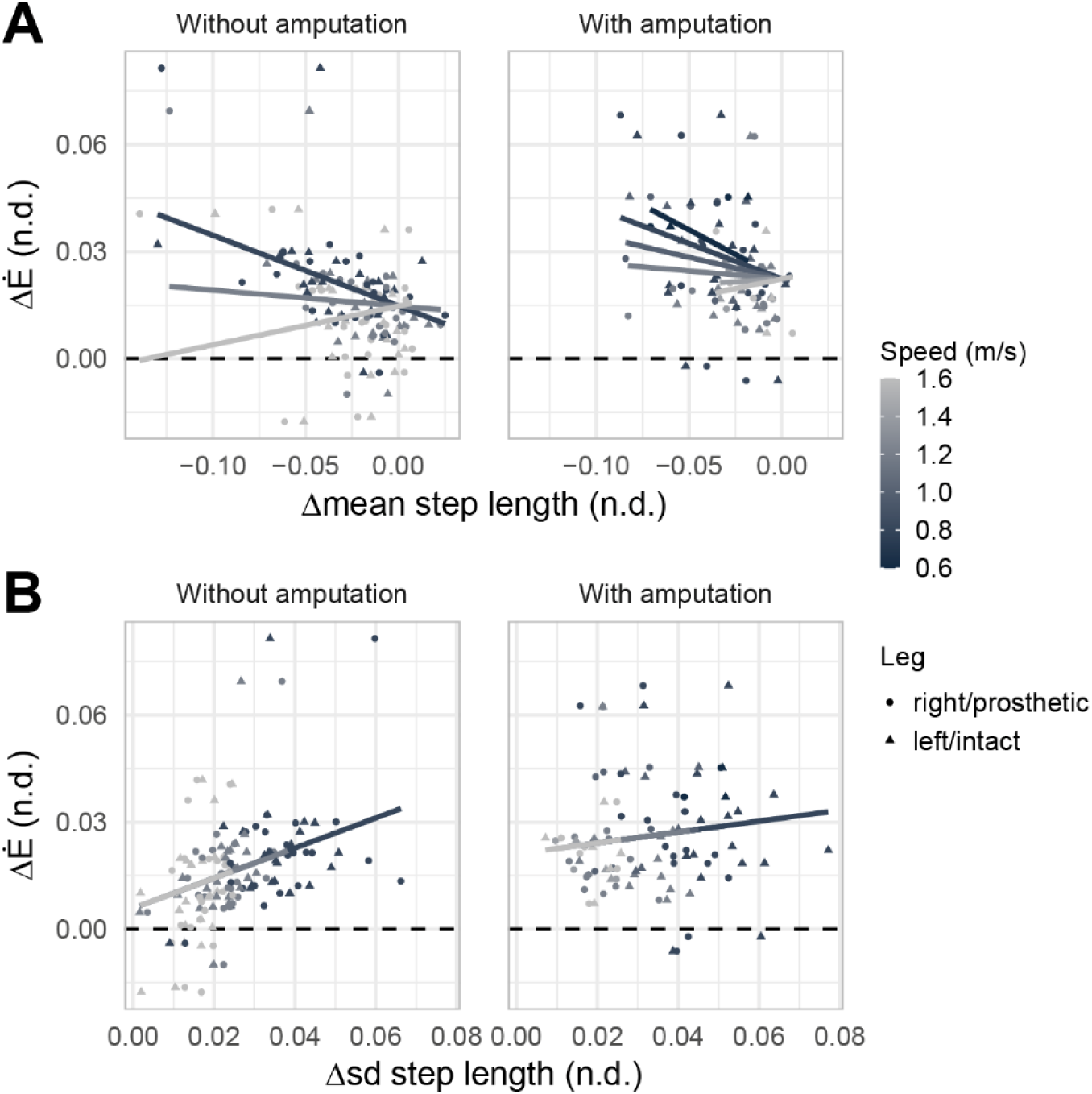
A. Change in metabolic rate due to perturbations against change in mean step length for subjects without (left) and with (right) a transtibial amputation. **B** Change in metabolic rate against change the standard deviation (sd) of step length for subjects without (left) and with (right) a transtibial amputation. Symbols indicate data points for both legs, a dot for the right or prosthetic leg, and a triangle for the left or intact leg, and lines represent the fixed effects of the fitted model. Darker grey symbols represent slower speeds, while lighter grey symbols denote faster walking speeds, as indicated by the legend.

## DISCUSSION

Individuals with a unilateral transtibial amputation consumed more metabolic energy to stabilize walking in the sagittal plane than individuals without an amputation. Across walking speeds, individuals with a transtibial amputation did not only consume 0.52 W/kg more metabolic energy during unperturbed walking but also increased their metabolic energy consumption by 0.24 W/kg more in response to random treadmill belt speed perturbations (with standard deviation of 0.13 m/s) than individuals without an amputation. Individuals with an amputation did not rely more on anticipatory reductions in step length than individuals without amputation but they increased step length variability in their intact leg more than in their prosthetic leg and more than individuals without an amputation. This suggests that individuals with a transtibial amputation compensate for the inability to use the ankle strategy when standing on the prosthetic limb by relying more on step length adjustments of the intact leg. Individuals with and without amputation who decreased mean step length more in response to perturbations also increased energy consumption more, but less at faster than slower walking speeds. The relationship between increases in metabolic energy and step length variability was weaker in individuals with an amputation than in individuals without an amputation. This might reflect the reduced redundancy in reactive strategies to stabilize walking, i.e. no ankle strategy on prosthetic side, in individuals with an amputation. Our findings suggest that the metabolic cost of stabilizing walking should be considered when seeking to reduce the metabolic cost of walking in individuals with a transtibial amputation.

### The higher metabolic cost of walking in individuals with an amputation might be due to the higher metabolic cost of stabilizing walking

We used perturbations to probe the energetic cost of stabilizing walking and found that individuals with an amputation increased their metabolic cost more in response to sagittal plane perturbations than individuals without an amputation. Even in the absence of external perturbations, active stabilization is required (6) and this has been shown to contribute to the metabolic cost of walking in both individuals without (6) and with (15) amputation. Hence, it is possible that the higher metabolic cost in individuals with versus without an amputation during unperturbed walking can be attributed to differences in how sagittal plane walking is stabilized. Two recent studies demonstrated that stabilization by either handrail support or through laterally stabilizing the pelvis by an external device reduced the metabolic cost of walking in individuals with a transtibial amputation, although this reduction was not compared to or was not different from the reduction in individuals without an amputation (15, 16). The lack of differences in reductions of metabolic energy consumption when stabilizing the pelvis contrasts with our findings. There are several possible reasons for these seemingly contrasting results. First, transtibial amputation might have a larger effect on sagittal than on frontal plane stabilization of walking. Whereas the loss of sensory information might affect stability across planes, frontal plane stabilization relies more on stepping strategies whereas sagittal plane stabilization also relies heavily on the ankle strategy – especially at low speeds – in individuals without an amputation (10, 32). As amputation disables the ankle strategy but not the stepping strategy on the amputated side, it might have a larger effect on sagittal plane than on frontal plane stabilization of walking. Second, external stabilization of the pelvis might induce larger gait adaptations in individuals with an amputation that are not triggered by the reduced need for stabilizing walking, which might have resulted in increases in the metabolic cost of walking that masked reductions in the metabolic cost of stabilizing walking. In our study, we probed the effect of transtibial amputation on the energetic cost of stabilizing walking through a perturbation paradigm that did not constrain body segment movement.

### The lower preferred walking speed in individuals with an amputation (33) might amplify the effect of alterations in metabolic cost of stabilizing walking

We found that the metabolic cost of stabilizing walking in the sagittal plane is higher at slow compared to fast walking speeds in both individuals without and with amputation. As individuals with an amputation have a lower preferred walking speed, they may thus not (only) consume more energy due to altered control of walking in the absence of sensory information from and direct control over the amputated limb but also because stabilizing walking requires more energy at their lower preferred walking speed. We studied the energy consumption for stabilizing walking per unit time (i.e., metabolic rate), which was higher at slow than fast walking speeds. Yet, energy consumption for stabilizing walking per unit travelled (i.e., cost of transport) is even larger at slow than at fast walking speeds as at a slower speed a shorter distance is travelled in the same unit time. Quantifying differences in metabolic cost of stabilizing walking in the sagittal plane between populations thus requires matched speeds or the ability to account for the effect of speed.

### Individuals with a transtibial amputation did not rely more on anticipatory adaptations in step length but relied more on reactive stepping of the intact leg to stabilize walking against perturbations

Most studies that investigate the effect of transtibial amputation on strategies to stabilize walking focus on strategies for frontal plane stability (22, 34) or challenge stability in the frontal plane (20, 21, 23, 24). Yet, the ankle strategy, which is missing in individuals with a transtibial amputation on the amputated side, is thought to contribute most to stabilizing walking in the sagittal plane (10). To our knowledge, we are the first to evaluate the effect of transtibial amputation on strategies for stabilizing walking in the sagittal plane. We found that subjects, both with and without an amputation, decreased their mean step length in response to perturbations, and more so at slow than at fast walking speeds. The lack of difference in adjustments in mean step length in response to sagittal plane perturbations between individuals without and with an amputation is consistent with the lack of differences in adjustments in step width in response to frontal plane perturbations (20, 21). Individuals with a transtibial amputation relied more on reactive stepping of the intact leg to stabilize walking against perturbations, especially at slower walking speeds. In contrast to individuals without an amputation who can rely on an ankle strategy, reactive stepping may be the primary strategy to stabilize walking in individuals with a transtibial amputation when standing on the prosthetic leg.

### Inter-subject variability in the metabolic cost to stabilize walking against sagittal plane perturbations is more strongly associated with step length variability in individuals without than with an amputation, which might reflect a decrease in the redundancy of strategies in individuals with an amputation

Individuals without an amputation can rely on both an ankle or stepping strategy on both sides, and those who rely more on a reactive stepping strategy seem to increase their metabolic cost more (10.8 W per cm increase in step length variability). In contrast, individuals with an amputation cannot rely on the ankle strategy on the amputated side and must therefore rely more on reactive stepping with their intact leg. This more stereotypical reliance on a stepping strategy might explain why the use of a stepping strategy explains little of the variability in metabolic cost to stabilize walking in individuals with an amputation (3.7 W per cm increase in step length variability). We cannot fully explain the large inter-subject variability in the metabolic cost of individuals with a transtibial amputation with our current outcome measures. It is likely that some of the variability is due to differences in muscle coordination to achieve similar kinematic strategies, and this should be further investigated.

### Between-subject variability in the energy needed to stabilize walking was high in individuals with a transtibial amputation and most of this variability remains unexplained by differences in kinematic strategy

The cause of amputation likely contributes to inter-subject variability in the energy needed to stabilize walking. It is well known that the energetic cost of walking is higher for vascular than for avascular amputation (35) and notably stabilizing walking by handrail support decreased the metabolic cost of walking on a treadmill in individuals where amputation was caused by a vascular disease (15) but not in individuals with an amputation due to other conditions. Vascular diseases might be associated with poor sensory function or poor physical fitness, which might reduce the ability to quickly react to perturbations of walking. Other factors that we did not measure but might affect balance control are prosthesis characteristics (e.g., stiffness (36)) and time since amputation (37).

### Whereas age may affect balance control (38), it is unlikely that differences in age between individuals without and with amputation influenced our main results

In both individuals without and with an amputation, fall risk increases with age, suggesting age-related declines in balance control (38, 39). It is unclear whether the metabolic cost of stabilizing walking is also affected by age. Since the average age in the group of subjects with a transtibial amputation was 17.3 years higher than in the group without an amputation, we performed a post hoc analysis to evaluate the effect of age (Supplementary material S5). This analysis did not reveal any effects of age on the metabolic cost of unperturbed walking or stabilizing walking against perturbations. The age of the subjects had a small effect on changes in step length variability in response to perturbations in individuals with an amputation. For every 20 years increase in age, step length variability on the intact leg decreased by 0.6 cm. Hence, differences in changes in step length variability in response to perturbations between individuals with versus without amputation might have been even higher if age had been matched.

### Since sagittal and frontal plane control of walking is coupled, sagittal plane perturbations also elicited changes in strategies to control frontal plane stability, which in turn might have energetic consequences

Therefore, we performed a post hoc analysis (Supplementary material S4) to evaluate whether adaptations in frontal plane control of walking also contributed to differences in metabolic energy consumption between groups. Both groups similarly increased step width (0.026 m at 0.8 m/s) and step width variability (0.003-0.004 m) in response to perturbations, and these adaptations were speed-dependent. The energy needed to stabilize walking against sagittal plane perturbations increased more in subjects in whom adaptations in mean step width and step width variability were larger (1333 W/m, p<0.001, and 4052 W/m, p=0.01 ;Table S5). In line with our observations for step length variability, inter-subject differences in step width variability explained less of the inter-subject differences in energy consumption in individuals with versus without a transtibial amputation (-3125 W/m, p=0.01; Table S5). A possible interpretation is that also in the frontal plane transtibial amputation might limit the available kinematic strategies, whereas it might increase variability in the underlying muscle coordination strategy.

### We reported normalized energetic rates to account for differences in length and mass between individuals, but limb loss affects body mass and this might have introduced a bias

In literature, the metabolic cost of walking is typically normalized by subject plus prosthesis mass (40) or by subject mass (41). Compared to studies in which the metabolic cost of walking was normalized to subject mass, we underestimated walking metabolic rate and, as a result, the change in the metabolic cost to stabilize walking. For example, assuming a prosthesis mass of 2.2-2.8 kg (42), normalizing metabolic rate by subject mass, would have increased the metabolic cost of stabilizing walking by up to 0.026-0.034 W/kg for an average subject (∼77 kg). Normalization to subject mass would thus have increased differences between groups but might not be a good way to capture what happens with amputation. To an individual, changes in absolute metabolic cost after amputation matter. Since a prosthesis is typically lighter than a biological limb, the normalized metabolic cost would increase after amputation even if there was no increase in absolute metabolic cost. Our normalization method might thus have overestimated alterations in the metabolic cost of stabilizing walking due to amputation. However, this effect is small with respect to the differences we observed. For example, if the intact lower leg of an average subject is 4.6 kg (6.1% of body mass (43)), and the combined mass of the residual limb (1/4 of the intact leg) and prosthesis (2.2-2.8 kg (42)) is 3.4-4.0 kg, then normalization to an equivalent body mass without amputation would result in normalized metabolic costs of stabilizing walking that are on average 0.007-0.014 W/kg lower. This is only 5.8% of the difference in metabolic cost between groups. Hence, the effect of different definitions of body mass on the metabolic cost for stabilizing walking is low, and choosing a different method for normalizing energy consumption to body mass is not expected to affect the main findings.

In conclusion, we found important differences between subjects with and without a unilateral transtibial amputation in strategies and metabolic cost for stabilizing walking in the sagittal plane. Individuals with a transtibial amputation consume more metabolic energy to stabilize walking against sagittal plane perturbations. As continuous control is also required for unperturbed walking, an increased metabolic cost of stabilizing walking might also explain part of the higher metabolic energy consumption during unperturbed walking. It is possible that we underestimated increases in the metabolic cost for stabilizing walking because the main reason for amputation in our sample was traumatic and increases in metabolic cost of walking are higher in individuals with a vascular amputation. In addition, the mobility of individuals with an amputation who participated in the study was better than the average mobility of individuals with a transtibial amputation. Subjects with a transtibial amputation relied more on reactive steps using their intact leg, especially at slow walking speeds, which likely is a compensation for the inability to use the ankle strategy in the prosthetic leg. While between-subject variability in the metabolic cost of stabilizing walking was partially explained by anticipatory and reactive adaptations in step length, a large amount of between-subject variability in the metabolic cost for stabilizing walking remained unexplained by step length adaptation. Thus, other factors that contribute to between-individual differences in the energy for stabilizing walking remain to be investigated. Our findings stress the importance of studying sagittal plane control of walking and should be considered when seeking to reduce the metabolic cost or to restore balance control of walking in individuals with a transtibial amputation.

## SUPPLEMENTARY MATERIAL

Supplementary text describing deviations from the preregistration document (S1), sample size estimation (S2), predicted values for main outcomes based on mixed-effect models (S3), the analysis of changes in step width and the relationship between changes in step width and the energy needed to stabilize walking (S4), the effect of age on the main outcomes of the study (S5), and a description of the elimination of gait events (S6) can be found at: https://doi.org/10.17605/OSF.IO/Z4PFA.

## DATA AVAILABILITY

Data and code for the statistical analysis of this study are openly available at Open Science Framework (DOI: https://doi.org/10.17605/OSF.IO/Z4PFA).

## ACKNOWLEDGMENTS

The authors would like to thank Orthopedie Van Haesendonck (OVH, Leuven, Belgium). They have played an instrumental role in recruiting subjects. Our special gratitude goes out to Bart Willems and Lien Geutjens. We would like to thank all individuals who participated in this study, as well as the colleagues who supported the measurements. The authors are especially grateful for the support they received from Simon Leeman, Stan Huyghe, Ben Legon, and Giel Houben throughout the experiment.

## GRANTS

The project was funded by an FWO fellowship (1SF7322N) received by WM.

## DISCLOSURES

The authors declare that the research was conducted in the absence of any commercial or financial relationships that could be construed as a potential conflict of interest.

## AUTHOR CONTRIBUTIONS

FDG, RR, MA, and WM developed the study concept and designed the experiments. After the design of the study, FDG, MA, and WM preregistered the study. WM acquired and analyzed the data. FDG and WM wrote the manuscript. FDG and WM interpreted the results, and FDG, RR, MA, and WM edited the manuscript. All authors contributed to the article and approved the submitted version.

## Notes

### Competing Interest Statement

The authors have declared no competing interest.

### Author Declarations

The experimental protocol was approved by the Ethics Committee Research UZ / KU Leuven (S69186) and pre-registered on Open Science Framework after collecting data from subjects without an amputation and the first subject with an amputation.

